# Acute Kidney Injury and Perioperative complications based on modality of nephrectomy for the treatment of kidney cancer

**DOI:** 10.1101/2024.09.24.24314317

**Authors:** Maulinkumar N. Patel, Deep Shah, Heli Patel, Edgar A. Jaimes

## Abstract

**Background:** Nephrectomy, either partial or radical, is a key treatment for kidney cancer. The surgical approach—open, laparoscopic, or robot-assisted—can influence perioperative outcomes. This study evaluates acute kidney injury (AKI) and perioperative complications across these modalities.

**Methods:** We conducted a retrospective analysis using the National Inpatient Sample (NIS) of 2020–2021 to examine 12,676 kidney cancer patients who underwent nephrectomy. Patients were stratified by surgical approach: open, laparoscopic, and robot-assisted. Primary outcomes included AKI incidence, length of stay, and intra- and post-operative complications. Multivariable regression models adjusted for confounding variables, including age, sex, race, Charlson Comorbidity Index (CCI), hospital region, and teaching status.

**Results:** Patients undergoing open nephrectomy had the highest AKI rates, particularly for radical nephrectomy (22.13% vs. 16.52% for robotic, p < 0.001). Similarly, patients treated with open surgery had longer hospital stays and higher rates of complications, including pulmonary (7.71% vs. 4.17%, p < 0.001) and vascular (12.11% vs. 4.01%, p < 0.001) in radical nephrectomy. Multivariate analysis confirmed the increased risk of AKI and complications in open nephrectomy, with robotic-assisted surgery demonstrating the best perioperative outcomes.

**Conclusion:** Open nephrectomy, whether partial or radical, is associated with higher AKI rates and perioperative complications compared to laparoscopic and robot-assisted surgeries. Robot-assisted nephrectomy, with its lower complication rates and shorter hospital stays, may be the preferred approach when feasible. Further efforts to expand access to minimally invasive techniques could improve outcomes for kidney cancer patients.

## Introduction

Kidney cancer is one of the few cancers where cytoreductive nephrectomy-surgical removal of the main tumor-has proven to be quite significant. Based on the TNM staging, anatomical complexity of the tumor, surgical expertise, and pre-existing frailty of the patient, either partial or radical nephrectomy has been used as a treatment modality. Partial nephrectomy (Nephron-sparing surgery) is now the treatment of choice in cases where a healthy tissue can be spared.^1^ Still, at non-academic hospitals, partial nephrectomy is underutilized in patients with kidney cancer.^2,3^ Open nephrectomies are now reserved only for those cases where minimally invasive techniques are unavailable. As complicated cases can be handled with a minimally invasive procedure, robot-assisted partial nephrectomy is becoming increasingly common.

In terms of the risk of adverse cardiovascular events following surgery, partial nephrectomy has been found to have a longer-lasting preventive impact than radical nephrectomy.^4,5^ Similarly, when comparing the modes of surgery, robot-assisted partial nephrectomy is associated with shorter hospital stays and lower conversion rates to open/radical nephrectomy.^6^ As patients treated with surgical approaches have long-term 10-year survival rates (85-96% cancer-specific^7^), peri-operative complications are important predictors for event-free survival. Most of the existing literature focuses on refining the surgical technique (open vs. laparoscopic vs. hybrid-robotic)^8^, long-term survival predictions^9^, and quality of life post-surgery.^10^

A previous study based on a Scandinavian national sample reported that open surgeries are associated with twice as many complications as minimally invasive surgeries (p<0.001).^11^ Similarly, a tertiary care study conducted in Saudi Arabia reported similar results with higher blood loss, longer hospital stay, and more operative time for the open partial nephrectomy compared to minimally invasive partial nephrectomy.^12^ On the contrary, a literature review examining the difference in perioperative outcomes of robotic radical nephrectomy vs. laparoscopic radical nephrectomy didn’t find any significant differences.^13^ However, no study, particularly in the United States, compared all the modalities against all possible outcomes. Through the retrospective analysis of the National Inpatient Sample (2020-2021)^14^, we intend to fill out the gap in the literature by identifying the incidence of peri-operative complications among different types of nephrectomies in patients with kidney cancers.

## Methods

### Data Source

We utilized discharge data from the National Inpatient Sample (NIS) 2020–2021 to identify kidney cancer patients who underwent partial nephrectomy (PN) or radical nephrectomy (RN).^14^ Patients were stratified based on the type of surgical approach: open, laparoscopic (LAP), or robot-assisted surgery (RAS). Our assessment focused on acute kidney injury, length of stay, and peri-operative complications of patients undergoing these procedures.

The NIS, part of the Healthcare Cost and Utilization Project (HCUP) family of databases sponsored by the Agency for Healthcare Research and Quality (AHRQ), is the largest publicly available all-payer inpatient healthcare database, estimating around 35 million hospitalizations annually to inform decision-making at the national level.^14^ All diagnoses and procedures were coded using the International Classification of Disease 10th revision Clinical Modification (ICD-10-CM) and the ICD 10th revision Procedure Coding System (ICD-10-PCS).

Adult patients (≥ 18 years) with kidney cancer (ICD-10-CM codes C641, C642, and C649) who underwent either partial (ICD-10-PCS codes 0TB00ZZ, 0TB10ZZ, 0TB04ZZ, 0TB14ZZ) or radical nephrectomy (0TT00ZZ, 0TT10ZZ, 0TT04ZZ, 0TT14ZZ) were included in this study. Bilateral nephrectomy patients (0TT20ZZ, 0TT24ZZ) were excluded. Patients were stratified based on the type of procedure – open, laparoscopic, and robotic. Laparoscopic approach patients with ICD-10-PCS code of robotic-assisted procedures (8E0W3CZ, 8E0W4CZ, 8E0W7CZ, 8E0W8CZ) were classified as robotic-assisted surgery.

### Definition of Variables for Analysis

The primary endpoints of this study comprised the length of hospital stay, the occurrence of acute kidney injury (AKI), and the incidence of intra-operative and post-operative complications. These complications include cardiac, pulmonary, vascular, gastrointestinal complications, bleeding, and infections, all of which were identified using ICD-10-CM codes based on established methodologies. The specific ICD-10 codes employed to identify these complications are detailed in Supplementary Table 1.

To account for the presence of comorbid conditions, the study utilized the Deyo modification of the Charlson Comorbidity Index (CCI), applying coding algorithms for ICD-10-CM codes as described by Quan et al.^15,16^ The covariates analyzed included patient characteristics such as age at the time of admission, sex, race, hospital teaching status, hospital location, and CCI score. Age was recorded in years, while sex was categorized as male or female. Race was classified into four categories: White, Black, Hispanic, and Other, the latter of which encompasses Asian, Pacific Islander, Native American, and other racial groups. Hospital teaching status was categorized into teaching and non-teaching hospitals. Hospital location was defined by the region in which the hospital was situated, specifically categorized into Midwest, Northeast, South, and West. The CCI score was stratified into four categories: 0, 1, 2, and ≥3, reflecting the severity of the patient’s comorbid conditions.

### Statistical Analysis

Descriptive analyses were first conducted to characterize the patient population and describe the primary outcomes. Categorical variables were summarized using frequencies and proportions, while continuous variables such as age and length of stay were summarized using median and interquartile range (IQR). For bivariate analysis, chi-square tests were applied to categorical variables, and Fisher’s exact test was used when the expected cell size was ≤5. Continuous variables were compared using the Kruskal-Wallis test.

In the second phase, univariable and multivariable analyses were performed to assess the associations between surgical approach and outcomes. Negative binomial regression models were employed to analyze the length of stay, and logistic regression models were used to evaluate peri-operative complications. These models were adjusted for potential confounders, including age at admission, biological sex, race, hospital teaching status, hospital region, and the Charlson Comorbidity Index (CCI).

Subgroup analyses were conducted within the partial and radical nephrectomy groups, stratified by the type of surgical procedure—open, laparoscopic, and robotic surgery. All analyses were performed separately for partial and radical nephrectomy to account for differences in surgical techniques and patient characteristics.

The analysis and reporting adhered to the National Inpatient Sample (NIS) reporting guidelines, including the stipulation that any cell counts less than eleven were reported as “<11.” All statistical analyses and graphics were generated using STATA v18.0.^17^. We used a significant level of p<0.05 for all the tests.

## Results

### Descriptive Statistics

In the dataset for the NIS 2020-2021, there were 12,676 patients with renal cancer who received either a partial (n=4,572) or radical (n=8,104) nephrectomy (see Table 1). The median age and gender distribution were uniform across all surgical approaches for both types of nephrectomies. The results revealed a significant racial disparity in the radical nephrectomy group (p < 0.001). A larger percentage of White patients underwent robotic-assisted (73.5%) and open (74.34%) procedures compared to laparoscopic-only (68.52%) operations. Furthermore, the study observed notable disparities in hospital teaching status and geographical region. Specifically, in the radical nephrectomy group, a greater proportion of patients in teaching hospitals and the South region opted for open surgery. The Charlson Comorbidity Index (CCI) indicated that most patients had a CCI score of 0 or 1, and significant differences were observed across surgical approaches in both the partial and radical nephrectomy groups (p < 0.001). In both cohorts undergoing partial and radical nephrectomy, the open surgical approach was found to have a higher percentage of patients with a Charlson Comorbidity Index (CCI) score of ≥3, as compared to the laparoscopic and robotic-assisted approaches. In particular, 17.89% of patients in the partial nephrectomy group and 31.34% in the radical nephrectomy group undergoing open surgery had a CCI score of ≥3.

### AKI, length of stay, and peri-operative complications in partial nephrectomy patients

Acute kidney injury (AKI) demonstrated a notable disparity among patients who received partial nephrectomy through various surgical approaches. The frequency of AKI was highest in individuals who underwent open surgery, with a percentage of 17.58%, followed by those who underwent laparoscopic procedures, with a percentage of 11.39%, and those who underwent robotic-assisted procedures, with a percentage of 8.12%. The length of stay was notably longer for individuals who underwent open surgery, at a median of 3 days (interquartile range: 2-5), than for those who underwent robotic-assisted or laparoscopic surgery, at a median of 2 days (interquartile range: 1-3) (p < 0.001). In terms of peri-operative complications, it was observed that intraoperative complications were more prevalent in open surgery (1.68%) than in laparoscopic (<2.78%) and robotic-assisted (0.65%) procedures (p = 0.01). Additionally, pulmonary complications were also more common in the open surgery group (8.32%) compared to laparoscopic (4.05%) and robotic-assisted (3.69%) procedures (p < 0.001). Additionally, cardiac complications were significantly higher in the open surgery group (9.26%) compared to the robotic-assisted group (6.82%) (p = 0.04). However, other complications, such as gastrointestinal, vascular, and postoperative bleeding, did not show statistically significant differences across the surgical approaches.

### Association between type of procedure and adverse outcomes in partial nephrectomy patients

After conducting a multivariable analysis of patients who underwent partial nephrectomy, it was found that the open surgical approach was associated with a significantly higher odds of acute kidney injury (AKI) (adjusted odds ratio [aOR] 2.21, 95% confidence interval [CI] 1.75–2.79) and an increased length of stay (adjusted relative risk [aRR] 1.76, 95% CI 1.68–1.85) compared to the robotic-assisted approach. The laparoscopic approach was also associated with a higher odds of AKI (aOR 1.64, 95% CI 1.08–2.49) and an increased length of stay (aRR 1.18, 95% CI 1.09–1.28) compared to the robotic-assisted approach. Furthermore, the open approach was linked to increased odds of pulmonary complications (aOR 2.14, 95% CI 1.56–2.94).

### AKI, length of stay and peri-operative complications in radical nephrectomy patients

In the group of patients who underwent radical nephrectomy, AKI was most commonly observed in those who had open surgery (22.13%), followed by those who had laparoscopic (14.90%) and robotic-assisted (16.52%) procedures (p < 0.001). Furthermore, the median duration of hospitalization was extended for individuals who underwent open surgery (3 days, IQR: 2-5) in contrast to those who had robotic-assisted or laparoscopic surgery (2 days, IQR: 1-3) (p < 0.001). Intraoperative complications were more prevalent in the open surgery group (3.1%) than in the laparoscopic (1.6%) and robotic-assisted (1.43%) groups (p < 0.001). Patients who underwent open surgery experienced more pulmonary complications (7.71%) than those who had laparoscopic (4.12%) or robotic-assisted (4.17%) procedures (p < 0.001). The frequency of vascular complications was highest in the open surgery group (12.11%), which was more than in the laparoscopic (2.96%) and robotic-assisted (4.01%) groups, showing a similar trend (p < 0.001). Furthermore, postoperative bleeding was more common in the open surgery group (1.07%) than in the laparoscopic (0.68%) and robotic-assisted (0.42%) groups (p = 0.01). Although cardiac and gastrointestinal complications were more prevalent in the open surgery group, there were no statistically significant differences between the groups.

### Association between type of procedure and adverse outcomes in radical nephrectomy **patients**

In the multivariate analysis of patients who underwent radical nephrectomy, the open surgical approach was linked to higher likelihood of developing acute kidney injury (AKI) (aOR 1.39, 95% CI 1.20–1.61) and an increased length of stay (RR 1.68, 95% CI 1.61–1.75) compared to the robotic-assisted approach, after accounting for potential confounding factors, including age at admission, sex, race, hospital teaching status, hospital region, and the Charlson Comorbidity Index. The open surgical approach was associated with a higher odd of intraoperative complications (aOR 2.23, 95% CI 1.52–3.28), cardiac complications (aOR 1.21, 95% CI 1.01– 1.43), pulmonary complications (aOR 1.85, 95% CI 1.45–2.36), vascular complications (aOR 3.27, 95% CI 2.60–4.12), and postoperative bleeding (aOR 2.58, 95% CI 1.34–4.96).

## Discussion

This study offers a thorough examination of the perioperative outcomes linked to different nephrectomy techniques, including partial and radical procedures, which are employed for treating kidney cancer. Using the National Inpatient Sample (NIS) database for the years 2020-2021, we aimed to determine the incidence of acute kidney injury (AKI), the length of hospital stay, and various intraoperative and postoperative complications associated with each surgical approach. Our results highlight substantial disparities in outcomes based on the type of nephrectomy and the surgical modality utilized.

### Key Findings and Clinical Implications

Our study findings show that open nephrectomy, whether partial or radical, has higher perioperative complications compared to laparoscopic and robot-assisted approaches. Notably, the open surgical method led to significantly higher rates of AKI, longer hospital stays, and a greater occurrence of pulmonary and vascular complications. For patients who undergo partial nephrectomy, the AKI rate was higher in the open surgery group (17.58%) compared to the laparoscopic (11.39%) and robot-assisted (8.12%) approaches. Furthermore, the length of hospital stay is notably longer in the open surgery group. These findings align with existing literature, which suggests that minimally invasive approaches, particularly robot-assisted surgery, are associated with better postoperative outcomes and shorter recovery times.^18,19^ In the cohort of patients who underwent radical nephrectomy, open surgery was associated with the highest rates of acute kidney injury (AKI) at 22.13% and longer hospital stays than laparoscopic and robotic-assisted surgeries. In addition, intraoperative complications and postoperative bleeding were more common in the open-surgery group. These findings emphasize the potential benefits of minimally invasive techniques, particularly robot-assisted surgery, in reducing the risk of adverse outcomes.

### Comparative Outcomes Between Surgical Modalities

The analysis of the three surgical methods showed that robotic nephrectomy performed better than both open and laparoscopic procedures in terms of decreasing the risk of acute kidney injury, shortening hospital stays, and reducing the likelihood of complications occurring during the procedure. The use of robot-assisted surgery for nephrectomy is becoming increasingly popular due to its precision and control, which may lead to improved outcomes. Our multivariate analysis supports this, as we found that after adjusting for potential confounding factors, the open surgical approach was associated with a significantly higher risk of developing AKI and a longer length of stay compared to the robot-assisted approach in both the partial and radical nephrectomy groups. These results are consistent with those of previous studies that have demonstrated the superiority of robot-assisted surgery in terms of perioperative safety and recovery.^20,21^

### Implications for Surgical Practice

The research indicates that robot-assisted nephrectomy should be given preference, particularly in situations where the patient’s health allows for minimally invasive surgery. Given the connection between open surgery and higher complication rates, surgeons should thoughtfully consider the potential risks and benefits of open nephrectomy, particularly for patients with significant underlying health conditions. In addition, our research emphasizes the necessity of ongoing advancements in minimally invasive surgical procedures and advocates for the broader implementation of these approaches in clinical practice. The literature has revealed an underutilization of partial nephrectomy, particularly in non-academic centers,^2,3,22,23^ indicating a possible area for enhancing patient care by providing the most effective and suitable surgical intervention.

### Limitations and Future Research

Although our study offers valuable insights into the outcomes associated with various nephrectomy modalities, it has limitations that should be considered. The study’s retrospective design and use of administrative data may introduce selection and reporting biases, potentially limiting the generalizability of the findings. However, this limitation is common to all previous analyses that rely on the National Inpatient Sample (NIS). ^24,25^ A potential shortcoming of current research is the scarcity of data on long-term outcomes, including quality-of-life measures following surgery. Future studies should focus on addressing this limitation by exploring the long-term complications associated with various nephrectomy methods and evaluating the influence of surgical expertise and institutional factors on patient outcomes. Furthermore, incorporating patient-reported outcomes in prospective studies would offer a more comprehensive perspective on the advantages and disadvantages of each surgical approach.

### Conclusion

In summary, this research emphasizes the benefits of minimally invasive surgical techniques, particularly robotic-assisted nephrectomy, in decreasing perioperative complications and enhancing recovery outcomes for patients with kidney cancer. The findings recommend the wider implementation of robot-assisted surgery and suggest that open nephrectomy should only be used when minimally invasive approaches are not suitable. Given the ongoing advancements in surgical technologies, it is crucial for healthcare providers to stay up to date with the latest innovations in delivering optimal care to their patients.

## Data Availability

All data produced in the present work are contained in the manuscript.

https://hcup-us.ahrq.gov/db/nation/nis/nisdbdocumentation.jsp

## Appendix

**Table 1:** Descriptive Statistics of Renal cancer patients who underwent partial or radical nephrectomy, stratified by surgical approach. (N= 12,676)

|  | Partial Nephrectomy |  |  |  | Radical Nephrectomy |  |  |  |
| --- | --- | --- | --- | --- | --- | --- | --- | --- |
|  | Robotic Assisted<br>(n=3,227) | Laparoscopic<br>(n=395) | Open<br>(n= 950) | p-value<sup>a</sup> | Robotic Assisted<br>(n=2,591) | Laparoscopic<br>(n=2,061) | Open<br>(n=3,452) | p-value<sup>a</sup> |
| **Age in years, median (IQR\*)** | 61 (52, 69) | 61 (52, 68) | 62 (52, 69) | 0.81 | 66 (57, 72) | 65 (56, 73) | 65 (56, 72) | 0.16 |
| **Gender** |  |  |  |  |  |  |  |  |
| **Males, n (%)** | 2,017 (62.52) | 251 (63.54) | 608 (64) | 0.68 | 1,651 (63.75) | 1,317 (63.93) | 2,256 (65.35) | 0.37 |
| **Race, n (%)** |  |  |  |  |  |  |  |  |
| **White** | 2,262 (71.88) | 269 (70.24) | 651 (69.63) | 0.36 | 1,855 (73.5) | 1,369 (68.52) | 2,489 (74.34) | **<0.001** |
| **Black** | 344 (10.93) | 46 (12.01) | 121 (12.94) |  | 239 (9.47) | 267 (13.36) | 341 (10.19) |  |
| **Hispanic** | 340 (10.80) | 50 (13.05) | 97 (10.37) |  | 258 (10.22) | 236 (11.81) | 338 (10.1) |  |
| **Other** | 201 (6.39) | 18 (4.70) | 66 (7.06) |  | 172 (6.81) | 126 (6.31) | 180 (5.37) |  |
| **Teaching Hospital status, n (%)** | 2,847 (88.22) | 349 (88.35) | 814 (85.68) | 0.21 | 2,202 (84.99) | 1,709 (82.92) | 2,790 (80.82) | **<0.01** |
| **Hospital region, n (%)** |  |  |  |  |  |  |  |  |
| **Northeast** | 737 (22.84) | 114 (28.86) | 186 (19.58) | **0.02** | 441 (17.02) | 353 (17.13) | 518 (15.01) | **<0.001** |
| **Midwest** | 666 (20.64) | 52 (13.16) | 180 (18.95) |  | 719 (27.75) | 346 (16.79) | 827 (23.96) |  |
| **South** | 1,152 (35.70) | 165 (41.77) | 413 (43.47) |  | 921 (35.55) | 913 (44.30) | 1,518 (43.97) |  |
| **West** | 672 (20.82) | 64 (16.20) | 171 (18) |  | 510 (19.68) | 449 (21.78) | 589 (17.06) |  |
| **Charlson Comorbidity Index Score, n (%)** |  |  |  |  |  |  |  |  |
| **0** | 1,621 (50.23) | 202(51.14) | 405(42.63) | **<0.001** | 1,030 (39.75) | 828 (40.17) | 1,267(36.70) | **<0.001** |
| **1** | 771(23.89) | 98(24.81) | 220(23.16) |  | 506 (19.53) | 466 (22.16) | 692 (20.05) |  |
| **2** | 394 (12.21) | 40 (10.13) | 155 (16.32) |  | 359 (13.86) | 275 (13.34) | 411 (11.91) |  |
| **≥3** | 441 (13.67) | 55 (13.92) | 170 (17.89) |  | 696 (26.86) | 492 (23.87) | 1,082(31.34) |  |

<sup>a</sup> Kruskal Wallis test, and Chi-square test
BOLD values are statistically significant (p <0.05)
\*IQR – Interquartile range

**Table 2:** Acute Kidney Injury, length of stay, and peri-operative complication rates after partial or radical nephrectomy among kidney cancer patients, stratified by surgical approach.

|  | Partial Nephrectomy |  |  |  | Radical Nephrectomy |  |  |  |
| --- | --- | --- | --- | --- | --- | --- | --- | --- |
|  | Robotic Assisted (n=3,227) | Laparoscopic (n=395) | Open (n= 950) | p-value<sup>a</sup> | Robotic Assisted (n=2,591) | Laparoscopic (n=2,061) | Open (n=3,452) | p-value<sup>a</sup> |
| **Acute Kidney Injury, n (%)** | 262 (8.12) | 45 (11.39) | 167 (17.58) | **<0.001** | 428 (16.52) | 307 (14.90) | 764 (22.13) | **<0.001** |
| **Length of stay in days, median (IQR\*)** | **2 (1, 3)** | **2 (1, 3)** | **3 (2, 5)** | **<0.001** | **2 (1, 3)** | **2 (1, 3)** | **3 (2, 5)** | **<0.001** |
| **Intraoperative complications, n (%)** | 21 (0.65) | <11 (<2.78) | 16 (1.68) | **0.01** | 37 (1.43) | 33 (1.6) | 107 (3.1) | **<0.001** |
| **Cardiac Complications, n (%)** | 220 (6.82) | 28 (7.09) | 88 (9.26) | **0.04** | 267 (10.3) | 243 (11.79) | 419 (12.14) | 0.07 |
| **Pulmonary Complications, n (%)** | 119 (3.69) | 16 (4.05) | 79 (8.32) | **<0.001** | 108 (4.17) | 85 (4.12) | 266 (7.71) | **<0.001** |
| **Gastrointestinal Complications, n (%)** | 221 (6.85) | 25 (6.33) | 51 (5.37) | 0.27 | 228 (8.80) | 159 (7.71) | 317 (9.18) | 0.18 |
| **Vascular Complications, n (%)** | 54 (1.67) | 11 (2.78) | 23 (2.42) | 0.14 | 104 (4.01) | 61 (2.96) | 418 (12.11) | **<0.001** |
| **Postoperative Bleeding, n (%)** | 20 (0.62) | <11 (<2.78) | <11 (<1.16) | 0.92 | 11 (0.42) | 14 (0.68) | 37 (1.07) | **0.01** |

<sup>a</sup> Kruskal Wallis test, Chi-square test, Fischer-exact test
BOLD values are statistically significant (p <0.05)
\*IQR- Interquartile range

**Table 3:** Univariable and Multivariable regression models addressing acute kidney injury, length of stay, and post-operative complications in 4,572 kidney cancer patients who underwent partial nephrectomy, stratified by surgical approach.

|  | Univariate |  |  | Multivariate* |  |  |
| --- | --- | --- | --- | --- | --- | --- |
|  | Robotic Assisted | Laparoscopic<br>RR/OR (95% CI) | Open<br>RR/OR (95% CI) | Robotic Assisted | Laparoscopic<br>RR/OR (95% CI) | Open<br>RR/OR (95% CI) |
| **Acute Kidney Injury** | Ref | 1.46 (0.99, 2.13) | **2.41 (1.93, 3.02)** | Ref | **1.64 (1.08, 2.49)** | **2.21 (1.75, 2.79)** |
| **Length of stay** | Ref | **1.17 (1.08, 1.26)** | **1.83 (1.75, 1.92)** | Ref | **1.18 (1.09, 1.28)** | **1.76 (1.68, 1.85)** |
| **Cardiac Complications** | Ref | 1.04 (0.70, 1.55) | **1.40 (1.07, 1.81)** | Ref | 1.03 (0.68, 1.55) | 1.30 (0.99, 1.70) |
| **Pulmonary Complications** | Ref | 1.10 (0.65, 1.88) | **2.37 (1.75, 3.20)** | Ref | 1.19 (0.70, 2.02) | **2.14 (1.56, 2.94)** |
| **Gastrointestinal Complications** | Ref | 0.92 (0.61, 1.40) | 0.77 (0.56, 1.07) | Ref | 0.94 (0.61, 1.45) | 0.77 (0.55, 1.07) |
| **Vascular Complications** | Ref | 1.68 (0.88, 3.24) | 1.46 (0.89, 2.38) | Ref | 1.77 (0.91, 3.43) | 1.30 (0.79, 2.14) |

\*Adjusted for age at admission, biological sex, race, hospital teaching status, hospital region, and CCI
RR- Rate ratio, OR – Odds ratio
Rate ratio is used for length of stay (using negative binomial regression)
BOLD values are statistically significant (p <0.05)

**Table 4:** Univariable and Multivariable regression models addressing acute kidney injury, length of stay, and peri-operative complications in 8,104 kidney cancer patients who underwent radical nephrectomy, stratified by surgical approach.

|  | Univariate |  |  | Multivariate* |  |  |
| --- | --- | --- | --- | --- | --- | --- |
|  | Robotic Assisted | Laparoscopic<br>RR/OR (95% CI) | Open<br>RR/OR (95% CI) | Robotic Assisted | Laparoscopic<br>RR/OR (95% CI) | Open<br>RR/OR (95% CI) |
| Acute Kidney Injury,<br>n (%) | Ref | 0.88 (0.75, 1.05) | **1.44 (1.25, 1.66)** | Ref | 0.91 (0.76, 1.08) | **1.39 (1.20, 1.61)** |
| Length of stay | Ref | **1.09 (1.04, 1.14)** | **1.72 (1.65, 1.79)** | Ref | **1.09 (1.04, 1.14)** | **1.68 (1.61, 1.75)** |
| Intraoperative<br>complications | Ref | 1.12 (0.70, 1.81) | **2.21 (1.51, 3.23)** | Ref | 1.16 (0.71, 1.88) | **2.23 (1.52, 3.28)** |
| Cardiac<br>Complications | Ref | 1.16 (0.97, 1.40) | **1.20 (1.02, 1.42)** | Ref | 1.21 (1.0, 1.47) | **1.21 (1.01, 1.43)** |
| Pulmonary<br>Complications | Ref | 0.99 (0.73, 1.34) | **1.92 (1.51, 2.44)** | Ref | 0.98 (0.72, 1.34) | **1.85 (1.45, 2.36)** |
| Gastrointestinal<br>Complications | Ref | 0.87 (0.69, 1.08) | 1.05 (0.88, 1.25) | Ref | 0.86 (0.69, 1.08) | 1.06 (0.88, 1.27) |
| Vascular<br>Complications | Ref | **0.73 (0.54, 0.99)** | **3.29 (2.63, 4.13)** | Ref | 0.77 (0.56, 1.05) | **3.27 (2.60, 4.12)** |
| Postoperative<br>Bleeding | Ref | 1.60 (0.73, 3.53) | **2.54 (1.33, 4.85)** | Ref | 1.50 (0.66, 3.40) | **2.58 (1.34, 4.96)** |

\*Adjusted for age at admission, biological sex, race, hospital teaching status, hospital region, and CCI
RR- Rate ratio, OR – Odds ratio
Rate ratio is used for length of stay (using negative binomial regression)
BOLD values are statistically significant (p <0.05)

